# Changing trends of excess self-protective behavior, and association with belief in prevention myths during the COVID-19 epidemic in China: A panel study

**DOI:** 10.1101/2020.05.18.20102434

**Authors:** Xiaozhao Yousef Yang, Sihui Peng, Tingzhong Yang, Ian R.H. Rockett

## Abstract

**Objective:** This prospective observational study examined changing trends of excess self-protective behavior (EPB), and its association with perceived risk, severity and belief in prevention myths during the Chinese COVID-19 epidemic.

**Methods:** The study employed a longitudinal design. Participants were recruited for an online panel survey from chat groups on social media platforms. Descriptive statistics and the CATMOD program were used for data analysis.

**Findings:** Participants numbered 150 for the linkable baseline survey and 102 for the final survey. There were 5 waves of interviews. The prevalence of participants perceiving a personal risk of contracting COVID-19, and severe consequences of the disease, was 18.6% and 25.5%, respectively. Their prevalence had declined to 4.9% and 17.6%, respectively, by the last observation point. The 5 selected EPBs also manifested a decreasing trend. Belief in COVID-19 prevention myths trended upwards. Perceived risk was positively associated with each EPB, and perceived severity with disinfection of clothes and hoarding of products. Myth adherence was positively associated with disinfection of clothes and both hand washing and sanitization.

**Conclusion:** This study yielded new information about EPB among the public during the COVID-19 epidemic. Policy and health education modifications are essential for minimizing the adverse health effects of subscribing to prevention myths.

## Introduction

Following the initial Wuhan outbreak in Hubei province, China, COVID-19 has rapidly diffused into a global pandemic [1]. Given the salience of human psychological and behavioral factors to disease prevention and mitigation, it is crucial to evaluate their role in propagating or impeding behavioral responses [2,3]. Many studies reported on mental and behavioral responses during an outbreak of an acute respiratory infection [4,5,6]. People quarantined at home or at another location may have experienced boredom, anger and loneliness, which in turn elicited a personal behavioral response[7,8].COVID-19 is a new disease, and the 2019 outbreak in Wuhan and elsewhere in China may have been stressful. An epidemic of a highly lethal disease can overwhelm people emotionally and physically, and induce strong mental and behavioral responses in both adults and children[5,9,10].

According to the Stimulus, Cognition and Response (SCR) model, various stimuli (S) affect internal states of people through cognition (C), which in turn elicits mental and behavioral responses (R)[8,11]. COVID-19 is a potent stimulus that plausibly induces people to perceive high risk of infection with potentially severe health consequences. Health belief theory proposes such perceptions may generate a behavioral response[8].Some studies found an association between strong risk and threat perceptions and excess mental and behavioral problems during outbreaks of the severe acute respiratory syndrome (SARS) and Ebola[12,13,14].A recent study revealed that individuals’ perceived severity of the COVID-19 epidemic was related to undesirable emotional and behavioral outcomes among the Chinese public[4].Perceived risk and severity of COVID-19 may also induce irrational beliefs about its prevention[15,16].Rational action theory proposes that all action is fundamentally ‘rational’ in terms of what the actor believes to be true, but in his or her practice comprises irrational as well as rational elements[17].Irrational beliefs are rigid, inaccurate and illogical, but are used defensively to process external events. Unlikely to find empirical support, these beliefs are self-defeating, unconditional and in conflict with reality[18].Many studies found that that the more irrational the belief, the more negative the health behaviours[16,18,19].Irrational beliefs commonly manifest in many social and health areas[8].

Survival is a biological imperative for humans, and self-protection is the common behavioral response for confronting a mortal crisis. In sudden major crises, coping can induce excess self-protective behavior (EPB). COVID-19’s evolution on the world scene has not been paralleled by any other communicable disease since the 1918 Spanish Flu pandemic; a catastrophic phenomenon that killed an estimated 50–100 million people globally. The dire threat to personal health and survival, posed by the COVID-19 pandemic, has quickly promoted perceptions of high risk for infection with potentially severe outcomes. In turn, these responses can be a strong stimulus for EPB. EPB may be disproportionate and not recommended as an effective response to the actual threat; thus, overburdening individuals and society and sparking diversion of scarce critical resources away from places where need for assistance is most acute[10].In cognitive science, EPB emanates from people’s fear and distorted view of the world[8]. Therefore, irrational beliefs or subscription to myths about COVID-19 prevention measures could stimulate pervasive EPB. EPB consumes a high degree of personal physical energy, and in the process diminishes disease immunity. Excessive protective measures can overstress healthcare facilities and other resources, and consequently exert a strong negative impact upon the economy and society as a whole[20].For example, panic buying of essential consumer items like toilet paper, first aid kits, bottled water, and hand sanitizer, in response to COVID-19, has led to global shortages and price gouging of consumer staples[20].This response impedes disease prevention and economic recovery. However, this issue has generated little discussion and debate[20], and no previous empirical studies have addressed EPB in relation to COVID-19 or other acute infectious respiratory diseases.

This study has three key objectives:

1. To examine levels of EPB by the Chinese amidst the COVID-19 epidemic.
2. To evaluate temporal trends in EPB during that epidemic.
3. To study the association between perceived risk and severity of COVID-19, and belief in related prevention myths, respectively, and EPB.

This study may yield information important in formulating policy and health education initiatives aimed at reducing EPB, with the goal of improving the design and targeting of effective interventions for preventing and mitigating COVID-19.

## Methods

### Study design

We conducted a prospective longitudinal observation study to examine temporal trends and changes in EPB, and its associations with selected perceptions and beliefs during the COVID-19 pandemic. Participants

Participants were recruited via a survey advertisement from social media groups on WeChat and Douban, two of the most popular social media platforms in China. Inclusion criteria were membership in a common community; being in the age group 20–60 years; having access to a Smartphone; knowing the Chinese language; and willing to participate in the panel study and provide follow-up information at the scheduled observation points. Participants were excluded if they refused to provide this information or had a medical condition that could limit or preclude their participation. Within the registration system, potential participants were screened to ascertain eligibility. Upon consent, participants received an electronic questionnaire and instructions on how to proceed. After reading the instructions, they were asked to provide an e-consent by tapping the “Confirmation and Authorization” button and then directed to the questionnaire. A special administrative WeChat group was established to manage the follow-up data collection, using a unique QR code for each respondent. The QR code was the vehicle not only for identifying unique participants but prohibiting non-participants from taking the survey. After scanning the QR code, survey participants could enter the investigation group without further preconditions.

This panel study analyzed five waves of data collected over a month: wave 1(5/Feb/2020), wave 2(12/Feb/2020), wave 3(19/Feb/2020), wave 4 (26/Feb/2020), and wave 5(4/March/2020). The entire observation period covered the peak and trough of the COVID-19 epidemic in China. Diagnosed patients respectively numbered 3,887, 2,015, 394, 433, and 133 at the time of each wave (National Health Commission of People’s Republic of China, 2020).

### Data Collection

An online survey was implemented on Wenjuanxing (https://www.wjx.cn), a survey service website similar to Qualtrics or Surveymonkey, but tailored to Chinese users. Each wave of the survey had a dedicated electronic questionnaire access link. The online questionnaire link was posted to the respondent group, centrally managed in a WeChat group, and accessible every Wednesday from 10:30 am to 4:30 pm. Data were collected from 9:00–11:00 am every Monday. Data collectors and facilitators were third-year doctoral students in a public health program. All responses were anonymous. The questionnaire took approximately 10 minutes to complete, and the same survey protocol was used for every wave of the survey to assure homogeneity of data administration and collection. This study was approved by the ethics committee of Zhejiang University. As appropriate, a token of appreciation, a total of 30 RMB was given to those participants who completed all 5 questionnaires.

### Measurement

In this study, basic individual demographic characteristics were tapped: age, gender, ethnicity, education level, marital status and occupation. Perceptions of risk and disease severity were respectively captured through the items “continual fear of infection by COVID-19”and “becoming infected by COVID-19 is a serious misfortune.”

This study addressed personal harboring of myths or irrational beliefs about effective COVID-19 prevention measures that were not founded upon reality and science. They reflected five common misconceptions that circulated during the COVID-19 epidemic in China: (1) Smokers are not susceptible to COVID-19, (2) Consuming alcohol can prevent the spread of the virus, (3) People should avoid people from Hubei province, where COVID-19 first manifested in China, in order to prevent contraction of the disease, (4) It is reasonable for employers to dismiss Hubei employees to prevent the spread of the novel virus, and (5) People who move away from an affected area should be deported back to their place of origin. Items were rated on a 5-point Likert-type scale, which ranged from 1 (strongly disagree) to 5 (strongly agree). Item scores were summed to attain a total score for belief in COVID-19 prevention myths. The higher that score, the greater the level of irrational prevention belief. The Cronbach’s α coefficient was 0.70, suggesting the questionnaire had acceptable reliability. Consistent with prior practices, a cutoff score of 15 or above signified strong acceptance or belief in prevention myths [8]. EPB was a common coping mechanism for confronting the COVID-2019 epidemic in China. Currently, there is no protocol for determining EPB in this situation. We identified them empirically from social norms perspective [8]. Through an online survey, study participants stated which of 16 types of behavior, aimed at preventing COVID-19, they would categorize as EPB. Complete responses were obtained from 116 participants. Where there was 80% agreement on a type among these participants, we preliminarily categorized it as EPB. These selections were then reviewed and approved by 12 health experts, and the 5 that received universal acceptance by the participants and experts were classified as EPB. They included in this study: (1) Clothing disinfected every one or two days (Disinfecting clothes), (2) Frequent hand washing beyond the regular washing before and after meals or after work (Washing hands), (3) Frequent use of a disinfectant when washing hands at home (Sanitizing hands), (4) Hoarding masks, alcohol and other forms of protective products (Hoarding products), and (5) Making arrangements for another or others to handle their family and occupational responsibilities if, and when, they contracted COVID-19 (Transferring responsibilities).

### Data analysis

All data were entered into a database using Microsoft Excel. They were then imported into SAS (9.3version) for the statistical analysis. Across survey waves, descriptive statistics were calculated for belief in prevention myths, perceived high risk for contracting the disease, perceived high severity of disease consequences, and EPB prevalence. The CATMOD program was used to conduct repeated measures analysis of variance to determine changing trends across the five observation points, and to examine the association between perceived disease risk and severity, and belief in prevention myths, respectively, with EPB using the method of weighted least squares[21].

## Results

One hundred-and-fifty participants were recruited at baseline. The baseline was linkable and there were three intermediate and a final observation point, with 102 participants available for analysis throughout; 99 came from 24 provinces located across China, differentiated by region. The remaining 3 were international.

Of the study sample, 61.8% were female and 93.1% were Han Chinese. The average age of participants was 39.1 years (SD: 12.5), 43.1%were never married, and 50.0% were married (Table 1). The prevalence of belief in COVID-19 prevention myths was higher among males than females (OR: 3.33) and increased with age (OR: 6.64, 7.31). Married people had a lower prevalence of high perceived risk (OR: 0.36), and the middle-aged (40–49 years) had a higher prevalence of perceived disease severity than comparison groups (OR: 1.85). Disinfection of clothes was less prevalent among females than males (OR: 0.34), and among professionals than people in other occupational groups (OR: 0.35). Hand washing was more prevalent among females than males (OR: 3.48) and less prevalent among the married than the never married (OR: 0.43). Hoarding products was more prevalent among females (OR: 2.46), the middle-aged (OR: 4.32), the least educated (OR: 0.32), and the married than their respective demographic counterparts (OR: 0.13).

**Table 1.** Sample characteristics and prevalence of prevention myth belief, perceived disease risk and severity, and excessive self-protective behavior.

| Characteristic | N | % | Myth belief(%) | OR(Adjusted OR) | Perceived risk(%) | OR(Adjusted OR) | Perceived severe (%) | OR(Adjusted OR) | Disinfecting clothes (%) | OR(Adjusted OR) | Washing hands (%) | OR(Adjusted OR) | Sanitizing hands (%) | OR(Adjusted OR) | Hoarding products (%) | OR(Adjusted OR) | Transferring responsibilities(%) | OR(Adjusted OR) |
| --- | --- | --- | --- | --- | --- | --- | --- | --- | --- | --- | --- | --- | --- | --- | --- | --- | --- | --- |
| *Sex* |  |  |  |  |  |  |  |  |  |  |  |  |  |  |  |  |  |  |
| male | 39 | 38.2 | 15.5 | 1.00 | 7.7 | 1.00 | 21.5 | 1.00 | 72.8 | 1.00 | 12.8 | 1.00 | 18.9 | 1.00 | 4.1 | 1.00 | 18.0 | 1.00 |
| female | 63 | 61.8 | 5.4 | 0.30**(0.14**) | 12.1 | 1.64 | 19.7 | 0.89 | 52.7 | 0.42**(0.34**) | 28.6 | 2.72**(3.48)** | 19.0 | 1.01 | 9.5 | 2.46** | 17.1 | 0.94 |
| *Age (years)* |  |  |  |  |  |  |  |  |  |  |  |  |  |  |  |  |  |  |
| <30 | 37 | 36.3 | 2.2 | 1.00 | 10.3 | 1.00 | 15.8 | 1.00 | 63.6 | 1.00 | 27.2 | 1.00 | 21.2 | 1.00 | 4.9 | 1.00 | 13.0 | 1.00 |
| 40-49 | 28 | 27.4 | 12.9 | 6.64**(8.25**) | 12.1 | 1.20 | 25.7 | 1.85* | 51.4 | 0.91 | 20.0 | 0.67 | 21.4 | 1.01 | 12.8 | 2.87**(4.32**) | 19.3 | 1.59 |
| 50+ | 37 | 36.3 | 14.0 | 7.31**(12.56**) | 9.1 | 0.87 | 21.0 | 1.42 | 56.4 | 0.74 | 19.9 | 0.66 | 15.0 | 0.66 | 5.9 | 1.22(3.21) | 20.4 | 1.71 |
| *Education* |  |  |  |  |  |  |  |  |  |  |  |  |  |  |  |  |  |  |
| High school or junior college | 50 | 49.0 | 13.6 | 1.00 | 10.4 | 1.00 | 20.0 | 1.00 | 59.6 | 1.00 | 22.0 | 1.00 | 18.0 | 1.00 | 10.0 | 1.00 | 20.0 | 1.00 |
| College and more | 52 | 51.0 | 5.4 | 0.36** | 10.3 | 0.99 | 20.7 | 1.05 | 61.2 | 1.07 | 23.1 | 1.06 | 20.0 | 1.14 | 5.1 | 0.48*(0.32**) | 15.1 | 0.71 |
| *Ethnicity* |  |  |  |  |  |  |  |  |  |  |  |  |  |  |  |  |  |  |
| Han | 95 | 93.1 | 9.9 | 1.00 | 11.0 | 1.00 | 20.0 | 1.00 | 60.5 | 1.00 | 22.6 | 1.00 | 19.0 | 1.00 | 7.6 | 1.00 | 16.7 | 1.00 |
| minority | 7 | 6.9 | 2.8 | 0.26 | 2.9 | 0.23 | 25.0 | 1.33 | 58.3 | 0.92 | 22.4 | 0.98 | 19.4 | 1.03 | 5.6 | 0.72 | 27.8 | 1.92 |
| *Marital status* |  |  |  |  |  |  |  |  |  |  |  |  |  |  |  |  |  |  |
| never married | 44 | 43.1 | 4.1 | 1.00 | 14.7 | 1.00 | 23.4 | 1.00 | 60.1 | 1.00 | 28.3 | 1.00 | 20.2 | 1.00 | 10.1 | 1.00 | 20.6 | 1.00 |
| married | 51 | 50.0 | 13.2 | 3.54** | 5.8 | 0.36** | 19.1 | 0.77 | 63.4 | 1.15 | 18.3 | 0.58*(0.43**) | 18.3 | 0.89 | 4.2 | 0.40**(0.13**) | 14.8 | 0.67 |
| Divorce or widowed | 7 | 6.9 | 14.7 | 3.87* | 4.0 | 1.20 | 11.4 | 0.42 | 40.0 | 0.44* | 20.0 | 0.42(0.41) | 17.1 | 0.82 | 8.32 | 1.49(0.32) | 17.1 | 0.80 |
| *Occupation* |  |  |  |  |  |  |  |  |  |  |  |  |  |  |  |  |  |  |
| manager | 34 | 33.3 | 11.0 | 1.00 | 9.9 | 1.00 | 23.6 | 1.00 | 67.0 | 1.00 | 28.8 | 1.00 | 19.8 | 1.00 | 7.7 | 1.00 | 20.3 | 1.00 |
| professional | 40 | 39.2 | 8.5 | 0.75 | 7.9 | 0.79 | 13.2 | 0.49 | 49.7 | 0.49**(0.35**) | 21.8 | 0.77 | 16.4 | 0.80 | 4.2 | 0.53 | 15.3 | 0.71 |
| Others | 28 | 27.5 | 8.6 | 0.77 | 14.4 | 1.53 | 25.9 | 1.13 | 66.2 | 0.96 | 20.0 | 0.73 | 21.6 | 1.12 | 11.5 | 1.56 | 16.5 | 0.78 |

\*p<0.05, \*\*P<0.01

All participants knew the disease had attained epidemic proportions and was highly contagious at the time of the first survey was implemented. The prevalence at baseline of the perceptions of high risk of contracting COVID-19 and disease severity was 18.6% and 25.5%, respectively, and declined to 4.9% and 17.6% by the last observation point—a statistically significant change. The prevalence of the five types of EPB showed a statistically significant downwards trend across the total observation period of this panel study. Simultaneously, there was a statistically significant upwards trend in belief in COVID-19 prevention myths (Table 2).

**Table 2.** Time change trend in prevention myth belief, perceived disease risk and severity, and excessive self-protective behavior.

| Survey Wave | Myth belief | Perceived risk | Perceived severity | Disinfecting clothes | Washing hands | Sanitizing hands | Hoarding products | Transferring responsibilities |
| --- | --- | --- | --- | --- | --- | --- | --- | --- |
| *Time1* | 5.9 | 18.6 | 25.5 | 66.8 | 28.4 | 24.4 | 10.8 | 23.5 |
| *Time2* | 8.8 | 12.7 | 19.6 | 62.8 | 22.5 | 18.5 | 7.9 | 14.7 |
| *Time3* | 9.8 | 6.8 | 20.6 | 62.7 | 25.5 | 20.2 | 7.8 | 15.7 |
| *Time4* | 11.8 | 8.0 | 18.6 | 41.2 | 20.6 | 19.3 | 2.9 | 13.7 |
| *Time5* | 10.8 | 4.9 | 17.6 | 44.4 | 15.7 | 17.7 | 7.8 | 19.2 |
| *Mean(SD)* | 9.4 | 10.4 | 20.4 | 60.4 | 22.6 | 19.0 | 7.4 | 17.5 |
| *Trend test($\beta, p$)* | 0.1412(0.0172)** | -0.1392(0.0177)** | -0.1192(0.0176)** | -0.0392(0.0178)* | -0.1149(0.0175)** | -0.1220(0.0176)** | -0.1451(0.0175)** | -0.1251(0.0174) |

Perceived high risk for contracting COVID-19 was positively associated with each type of selected EPB, and perception that the disease had severe consequences was positively associated with disinfection of clothes and hoarding of products. Belief in the disease prevention myths was positively associated with disinfection of clothes and both hand washing and sanitization (Table 3).

**Table 3.** Association of perceived disease risk and severity and prevention myth belief with excessive self-protective behavior.

| Group | N(%) | Disinfecting clothes |  | Washing hands |  | Sanitizing hands |  | Hoarding material |  | Transferring responsibilities |  |
| --- | --- | --- | --- | --- | --- | --- | --- | --- | --- | --- | --- |
|  |  | Prevalence (%) | Parameter estimate(SE) | Prevalence (%) | Parameter estimate(SE) | Prevalence (%) | Parameter estimate(SE) | Prevalence (%) | Parameter estimate(SE) | Prevalence (%) | Parameter estimate(SE) |
| *Perceived risk* |  |  | 0.2500(0.0127)** |  | 0.0608(0.0101)** |  | 0.0413(0.0108)** |  | 0.02677(0.00775)** |  | 0.0353(0.0104)** |
| no | 457(89.6) | 60.0 |  | 19.3 |  | 18.6 |  | 5.3 |  | 16.4 |  |
| yes | 53(10.4) | 62.3 |  | 50.9 |  | 22.7 |  | 26.4 |  | 26.4 |  |
| *Perceived severity* |  |  | 0.2000(0.0130)** |  | 0.0108(0.0125) |  | -0.00686(0.0124) |  | 0.06470(0.00988)** |  | 0.0147(0.0117) |
| no | 406(79.6) | 56.9 |  | 21.4 |  | 19.1 |  | 5.4 |  | 15.8 |  |
| yes | 104(20.4) | 74.0 |  | 26.9 |  | 19.0 |  | 15.4 |  | 24.0 |  |
| *Myth belief* |  |  | 0.2599(0.2599)** |  | 0.0657(0.0117)** |  | 0.0489(0.0107)** |  | 0.00980(0.00854) |  | 0.0402(0.0022) |
| low | 462(90.6) | 60.1 |  | 16.6 |  | 18.6 |  | 7.1 |  | 16.5 |  |
| high | 48(9.4) | 62.5 |  | 25.8 |  | 22.9 |  | 10.4 |  | 27.1 |  |

## Discussion

At baseline, this study found that 18.6% and 25.5% participants, respectively, believed they were at high risk of contracting COVID-19, and that this disease seriously threatened their health. Turning to previous studies, one found approximately 10–30% of the general public were very worried or moderately worried about the possibility of contracting influenza during an outbreak [22]. Another reported that during the period February 1^st^ through 10^th^, when disease cases were increasing dramatically, 15.3% of Shanghai and Wuhan residents perceived COVID-19 as a very serious disease [23]. Our research commenced at the peak of the epidemic, and one-fifth to one-quarter of study participants perceived the disease was very serious and their risk of contraction was high.

COVID-19 is a new disease, and the epidemic may profoundly impact people mentally and behaviorally. Viewed as a stimulus, this disease can be overwhelming and elicit strong mental and behavioral responses. Many studies have found that COVID-19, now a global pandemic, generates negative mental and behavioral outcomes[9,10] that may include inappropriate health-protective and help-seeking behaviours[20].

Addressing a gap in the literature, this Chinese study found changing temporal trends in perceived high risk of contracting COVID-19, perception of severe adverse consequences of the disease, belief in prevention myths about the disease, and EPB during the epidemic. Prevalence of perceived risk and severity declined over the observation period, a trend consistent with the decline in the actual risk of infection. The numbers of new patients in China diagnosed with COVID-19 across the 5 observation points were 3,887, 2,015, 394, 433 and 133, respectively[24].As COVID-19 was reaching epidemic proportions, there were increases in the prevalence of perceptions of personal risk for contracting the disease and the severity of its health consequences. The prevalence of the five types of EPB, highlighted in this study, showed a statistically significant decreasing trend over the observation period. EPB is typically considered a behavioral overreaction in China, and increases with perceived disease risk and severity. Plausibly then, high perceived risk and severity induce EPB. Contrasting with our findings about the perception data, belief in COVID-19 prevention myths manifested a statistically significant increasing trend over the observation period. The explanation may inhere in an “energy consumption” mechanism [25].People likely functioned rationally as they mobilized all of their physical and mental energy to cope with COVID-19 at the beginning of the epidemic. However, as time passed, such energy waned and rational thinking diminished as belief in prevention myths became more common. This information is useful for formulating prevention policy and educational programming.

The risk of disease or injury and the severity of outcomes are crucial themes in individual health behavior. This study provided new evidence that perceived risk of contracting COVID-19 and perception of the severity of its consequences were both positively associated with several types of EPB, findings generally compatible with those from some other studies[3,4,12].We found a negative association between belief in prevention myths and some of the constituents of EPB. Affirmation for our findings, other investigators also found a relationship between such a belief and negative health behaviour[16.18].

EPB transcends normal self-protective behaviors, with special significance from a disease prevention perspective. For effective prevention, it is necessary both to avoid inadequate prevention measures that increase the likelihood of a disease epidemic, and to avoid excessive activities that waste personal and social resources. These two scenarios may vary across cultures. Inadequate prevention may be a prominent problem in Western culture and excessive prevention in Eastern culture[8]. A related dimension of such cultural variance is societal “rigidity” versus “porousness” [26].“Rigid” cultures, such as those of Singapore, Japan, and China, have strict social norms and punishment for deviance, whereas “porous” cultures, such as those characterizing the U.S., Italy, and Brazil, reflect weaker social norms and greater permissiveness[27].“Qǐ rén yōu tiān” from ancient China is a tale about a person who worried every day the sky would collapse. This study found that in the COVID-19 epidemic there was pervasive over-prevention among members of the public. Excessive prevention consumes too much personal energy and societal resources, and hence impedes disease control and economic recovery. Government and society at large must give this issue more attention. Reforms in health policy and health education will be essential for minimizing the adverse effects of belief in prevention myths and associated deleterious behavior.

There are two study limitations. First, our sample size is small. Nevertheless, the sample originated from 24 provinces covering diverse regions and a wide array of demographic characteristics. On the other hand, sample attrition may introduce a “cluster” bias since many longitudinal studies likely over-represent some of these characteristics, such as high educational attainment. A more sophisticated design and representative sample would be necessary to resolve this problem. A secondly limitation in this study is the lack of a clear definition of EPB. We operationalized this concept through empirically identifying 5 constituents from a social norms perspective.

Operationalization of the concept of belief in prevention myths may so be thought as an external criterion for measuring the validity of EPB [8,16,19]. Belief in prevention myths was significantly associated with 3 of the 5 types of EPB we utilized in this study. This finding enhances the validity of our measure of EPB. The concept and its operationalization requires further research

This study provides new information on the relationship between belief in COVID-19 prevention myths and EPB among the Chinese public, in the context of perceived risk of contracting the disease and perception of the severity of its consequences during the epidemic. As the virus spreads relentlessly around the globe, our findings could guide similar research outside China in less and more developed countries. They harbor important implications for understanding and decreasing EPB, as appropriate, during this new global pandemic.

## Data Availability

Data sharing not applicable to this article because the datasets we used belongs to our Centre for Tobacco Control Research Zhejiang University School of Medicine. Please contact corresponding author for data requests.

## Declaration of interests

The authors declare that they have no competing interests

## Acknowledgements

This study is partly supported by the National Nature Science Foundation of China (Major Project 71490733).

## Contributors

XYY and IRHR contributed to interpreting results, drafting and revising the manuscript. TZY conceived the study, conducted analyses, and led the writing. SHP conducted the literature search and data collection.

## Notes

### Competing Interest Statement

The authors have declared no competing interest.

### Funding Statement

This study is supported by the National Nature Science Foundation of China (Major Project 71490733).

